# Elective Surgery Before, During and After the COVID-19 Pandemic in England 2015 - 2022: A Database Study

**DOI:** 10.1101/2023.01.20.23284826

**Authors:** Sandra Remsing, Katharine Reeves, Felicity Evison, Dion Morton, Peter Chilton, Paul Bird, Samuel Watson, Kamlesh Khunti, Richard Lilford

## Abstract

**Objective:** To track elective surgery activity before, during and after the COVID-19 pandemic in England. To examine for hypothesised differences in use of independent vs NHS hospitals, and more urgent vs less urgent operations over the pre- and post-COVID time windows.

**Design:** We extracted data from the Hospital Episodes Statistics database from 1^st^ April 2015 to 30^th^ April 2022. This database contains all emergency and elective patient admissions, outpatient appointments and A&E attendances funded by the NHS in England.

**Setting:** NHS and Independent hospitals in England.

**Participants:** Adult patients (over 18 years) admitted for elective surgery between April 2015 and April 2022, who were classified as being in priority groups 3 or 4.

**Main Outcomes:** Total operations, operations by hospital type, and NHS England priority ranking.

**Results:** The data show that there was a large reduction in the number of elective operations during lockdown with incomplete recovery thereafter. Also the proportion of more urgent surgeries and surgeries in independent hospitals increased in the post-COVID vs pre-COVID time windows.

**Conclusion:** Under conditions of high-demand, higher value elective surgery procedures are awarded increasing priority and the Independent sector bears a larger share of the load.

## BACKGROUND

Waiting lists for planned surgery have increased rapidly during the COVID pandemic. As of July 2022, 377,689 people had been waiting more than a year for elective surgery in England.(1) In this study we examine the effect of COVID on access to planned surgery. We examined access over the COVID and post-COVID time windows with respect to the historic (pre-COVID) data.

Our purpose is to investigate the following questions:

1. What are the time-trends in operations before, during and after the COVID pandemic? Ideally, the post-COVID numbers would be higher than pre-COVID to compensate for reductions over the COVID period. However, there are many reasons why operation rates may not recover, despite increased demand in the post-COVID time window. These reasons include ongoing staff shortages and possible demoralisation/ exhaustion among the workforce.
2. Does the proportion of NHS-funded elective surgery shift towards the independent sector under high demand? Since operations on NHS-funded patients take place in independent providers as well as NHS owned hospitals, we hypothesised that the independent sector would act as an over-flow provider for the NHS in times of stress and increasing waiting lists. In that case, we would expect to observe higher proportions of elective surgeries in independent providers compared to NHS providers in the post-COVID time window.
3. Under high demand, does the service discriminate in favour of higher value / more urgent procedures? Elective surgeries are classified into four priority groups by NHS England’s Surgical Prioritisation Criteria, described in Supplementary Table S1.(2) In this paper, we focus on priority groups 3 and 4. If the service is prioritising effectively, then priority should be afforded to the higher priority operations post-COVID. This should show up as higher proportions of the more urgent category in the post-COVID time window compared to the pre-COVID time window.

There have also been reports of discrimination against some groups in access to elective surgery.(3) We therefore record differences in proportions of surgery across socio-economic and ethnic groups. However, this data has to be interpreted with caution for reasons given in the discussion. We also examine trends in surgery rates and socioeconomic group with the caveat that we describe below. For completeness we also look at outcomes of surgery, but again causal inferences must be treated with caution.

## METHODS

### Study design

The aim of this study is to examine the effect of COVID on access to planned NHS-funded surgery overall and at NHS vs independent hospitals and in Priority groups 3 and 4. We do this by plotting the monthly number of procedures before, during and after the COVID lockdowns and recording the percentages in different groups over these three time windows. We report the study in accordance with STROBE criteria.(4)

### Setting

The data used for this study were extracted from the Hospital Episodes Statistics (HES) database that contains all emergency and elective patient admissions, outpatient appointments and A&E attendances funded by the NHS in England. All procedures funded by the NHS were included, irrespective of whether they took place in NHS or independent hospitals.

### Participants

Admissions for elective surgery (spells) were included if they met the following criteria:

- One of the operation codes from Priority group 3 or 4 listed in Supplementary Table S2 in primary position.
- Operation date between 1^st^ April 2015 and 30^th^ April 2022.
- Not a repeat of a particular operation for a patient.
- Patient age on admission between 18 and 120.
- Admission method is elective (not emergency).

Patients with unknown sex or age, and patients with inconsistent data (e.g. date of death preceding operation date) were excluded.

### Variables

Information extracted about each surgical procedure is itemised in Table 1.

**Table 1:**
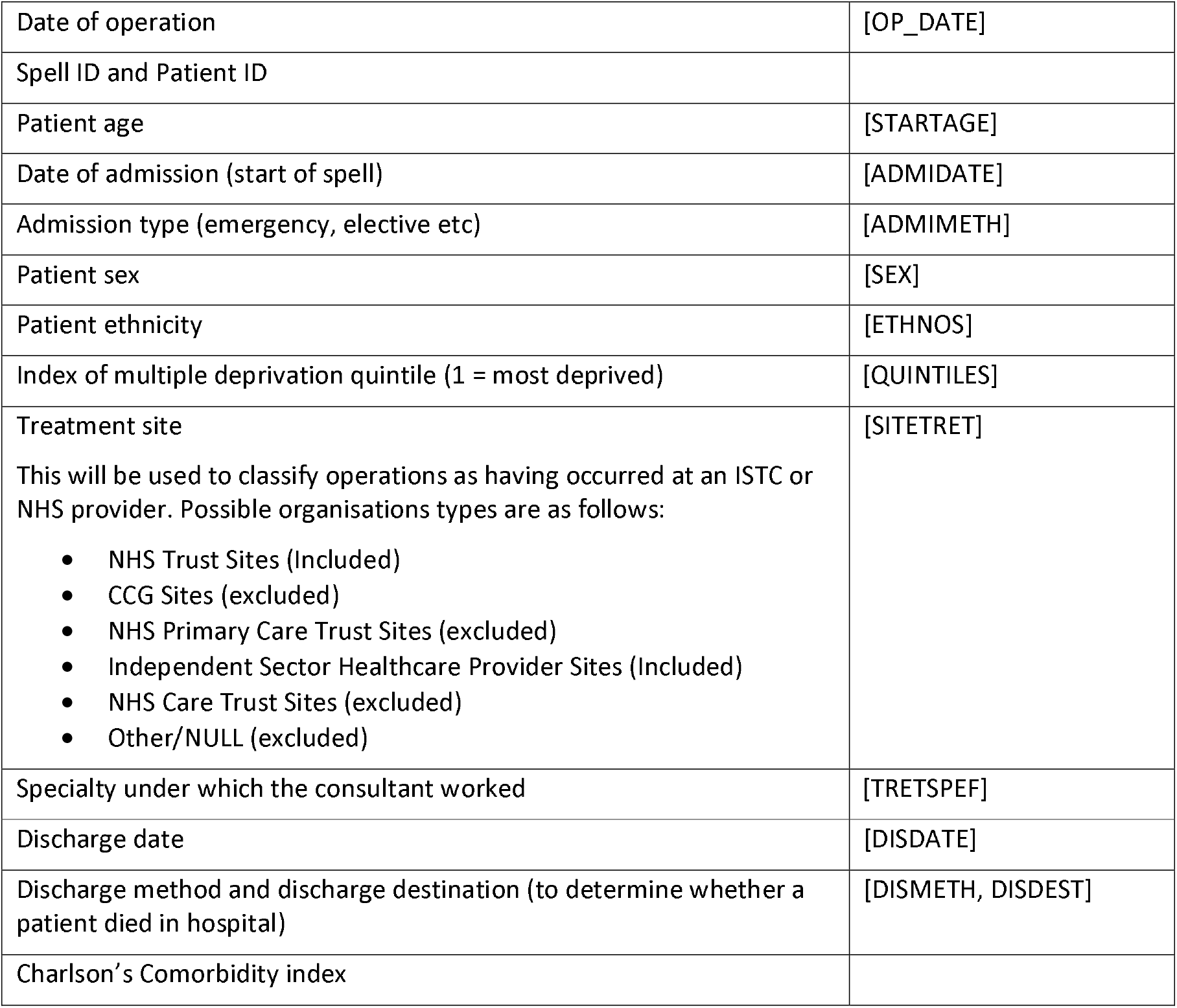
Data extracted for each operation

### Data sources

Data were extracted from the HES database. Our study was based on adult patients (aged ≥18 years old) with selected operation codes. We extracted data over three time windows: April 2015 – March 2000 (pre-COVID); April 2000 – March 2021 (COVID); and April 2021 –April 2022 (post-COVID).

We identified the procedure codes for the most frequently performed procedures from each of these two categories. The list of procedures used in this study, along with their OPCS4 codes, are given in Supplementary Table S2.

### Bias

In this study, we focus on the most frequently performed procedures from Priority groups 3 and 4. These procedures may not be representative of all procedures in these groups, but nevertheless provide a picture of the most common operations. We explore the issue of bias further in the discussion.

### Study size

In this study, we have included all procedures that satisfy the inclusion criteria. For Priority 3, there were 3,378,611 procedures and for Priority 4, there were 607,259 procedures.

### Statistical methods

For groups of patients treated at NHS sites and ISHP sites, we obtained:

1) Number of surgeries across the above three time-windows

2) Types of surgeries across the three time-windows.

Since the time-windows are of unequal duration, we made all comparisons per unit of time and include the monthly mean number of operations. We plotted the results across the time windows to allow visual inspection and describe the overall trends in operations.

We describe differences between pre and post-COVID proportions of surgeries by ethnic and socioeconomic group. However, it would not be appropriate to test the statistical significance of the difference between these proportions as we do not have data on the underlying proportions in the population.

In addition, we describe, across time-windows the ages of patients undergoing elective surgery, the comorbidity groups (using Charlson comorbidity scores), 90-day death rates and 90-day emergency readmission rates.

### Data access and cleaning methods

Investigators at University Hospitals Birmingham (UHB) have access to pseudonymised, patient-level HES data for the purposes of this study. Data were cleaned using the inclusion criteria listed above and aggregated for reporting purposes. This observational study was registered with the local Clinical Audit Department (Clinical Audit Registration and Management System number 17421). Data were used in line with the data sharing agreement with NHS Digital.

## RESULTS

### Participants

The disbursement of participants is shown in Figure 1.

**Figure 1:**
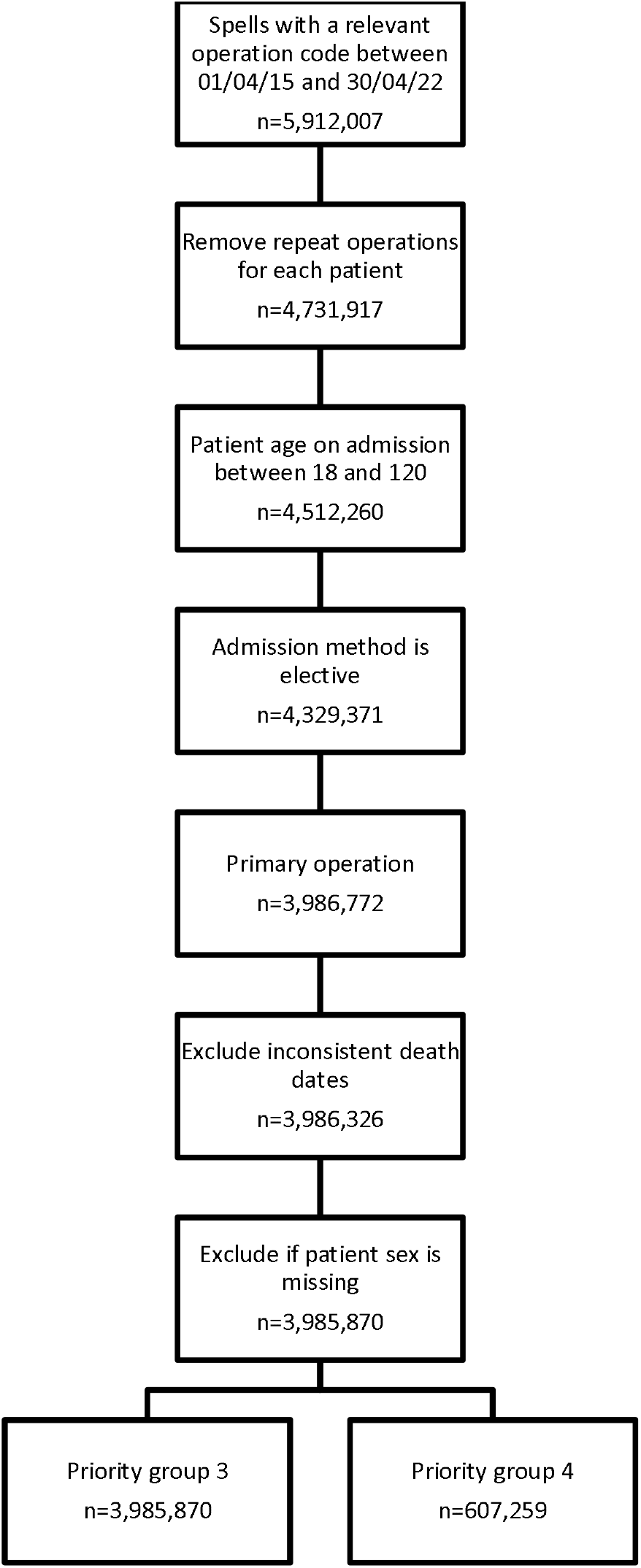
Disbursement of participants.

We identified 2,624,440 Priority 3 surgeries in time-window 1; 238,990 in time-window 2; and 515,181 in time-window 3 (Table 2). For Priority 4, the corresponding figures are 488,502; 41,886 and 76,871 (Table 3). Counts of operations performed over time can be visualised in Figure 2.

**Table 2:** Total operations and outcomes for Priority 3

| Priority 3 |  |  |  |  |  |  |
| --- | --- | --- | --- | --- | --- | --- |
|  | Pre-COVID<br>Apr 15 - Mar 19 |  | COVID<br>Apr 20 - Mar 21 |  | Post-COVID<br>Apr 21 - Apr 22 |  |
|  | Total | Monthly mean | Total | Monthly mean | Total | Monthly mean |
| Operations (total = 3,378,611) | 2,624,440 | 54,676 | 238,990 | 19,916 | 515,181 | 39,629 |
| **Provider** | n | % | n | % | n | % |
| NHS | 2,073,273 | 79.00% | 166,022 | 69.51% | 323,686 | 62.92% |
| ISHP | 551,100 | 21.00% | 72,829 | 30.49% | 190,781 | 37.08% |
| **Sex** | n | % | n | % | n | % |
| Male | 1,085,864 | 41.38% | 98,229 | 41.10% | 214,053 | 41.55% |
| Female | 1,538,576 | 58.62% | 140,761 | 58.90% | 301,128 | 58.45% |
| **Comorbidities** | n | % | n | % | n | % |
| 0 | 1,689,054 | 64.36% | 161,375 | 67.52% | 339,741 | 65.95% |
| 1-3 | 216,807 | 8.26% | 17,013 | 7.12% | 38,317 | 7.44% |
| 4+ | 718,579 | 27.38% | 60,602 | 25.36% | 137,123 | 26.62% |
| **Deaths from any cause (within 90 days)** | n | % | n | % | n | % |
| Total | 9,919 | 0.38% | 1,069 | 0.45% | 1,959 | 0.38% |
| **Emergency readmissions (within 90 days)** | n | % | n | % | n | % |
| Total | 141,541 | 5.39% | 12,566 | 5.26% | 27,015 | 5.24% |
| **Age (years)** |  |  |  |  |  |  |
| Median (IQR) | 71(60-79) |  | 71(60-78) |  | 72(62-79) |  |
| **LOS (days)** |  |  |  |  |  |  |
| Median (IQR) | 0(0-2) |  | 0(0-1) |  | 0(0-1) |  |
| **Ethnicity (n, %)** | n | % | n | % | n | % |
| Black | 45,435 | 1.73% | 3,640 | 1.52% | 8,616 | 1.67% |
| Mixed/other | 40,063 | 1.53% | 5,109 | 2.14% | 8,814 | 1.71% |
| South Asian | 102,939 | 3.92% | 8,093 | 3.39% | 19,048 | 3.70% |
| Unknown | 407,384 | 15.52% | 58,468 | 24.46% | 133,061 | 25.83% |
| White | 2,028,619 | 77.30% | 163,680 | 68.49% | 345,642 | 67.09% |
| **Deprivation (n, %)** | n | % | n | % | n | % |
| Quintile 1 (most deprived) | 439,294 | 16.74% | 38,948 | 16.30% | 83,840 | 16.27% |
| Quintile 2 | 491,645 | 18.73% | 43,765 | 18.31% | 94,246 | 18.29% |
| Quintile 3 | 550,714 | 20.98% | 49,684 | 20.79% | 107,813 | 20.93% |
| Quintile 4 | 568,106 | 21.65% | 52,635 | 22.02% | 113,327 | 22.00% |
| Quintile 5 (most affluent) | 557,580 | 21.25% | 52,488 | 21.96% | 112,796 | 21.89% |
| Unknown | 17,101 | 0.65% | 1,470 | 0.62% | 3,159 | 0.61% |
| **Operation** | n | % | n | % | n | % |
| Arthroplasty of joint | 65,990 | 2.51% | 4,741 | 1.98% | 10,815 | 2.10% |
| Endoscopic resection of prostate | 56,398 | 2.15% | 5,277 | 2.21% | 8,554 | 1.66% |
| Hernia repair | 121,966 | 4.65% | 8,285 | 3.47% | 18,780 | 3.65% |
| Hip replacement | 325,739 | 12.41% | 23,478 | 9.82% | 57,876 | 11.23% |
| Knee replacement | 343,144 | 13.07% | 19,642 | 8.22% | 52,359 | 10.16% |
| Lens replacement (cataracts) | 1,307,179 | 49.81% | 133,333 | 55.79% | 293,702 | 57.01% |
| Total abdominal hysterectomy | 120,171 | 4.58% | 15,474 | 6.47% | 23,080 | 4.48% |
| Total cholecystectomy | 283,853 | 10.82% | 28,760 | 12.03% | 50,015 | 9.71% |

**Table 3:**
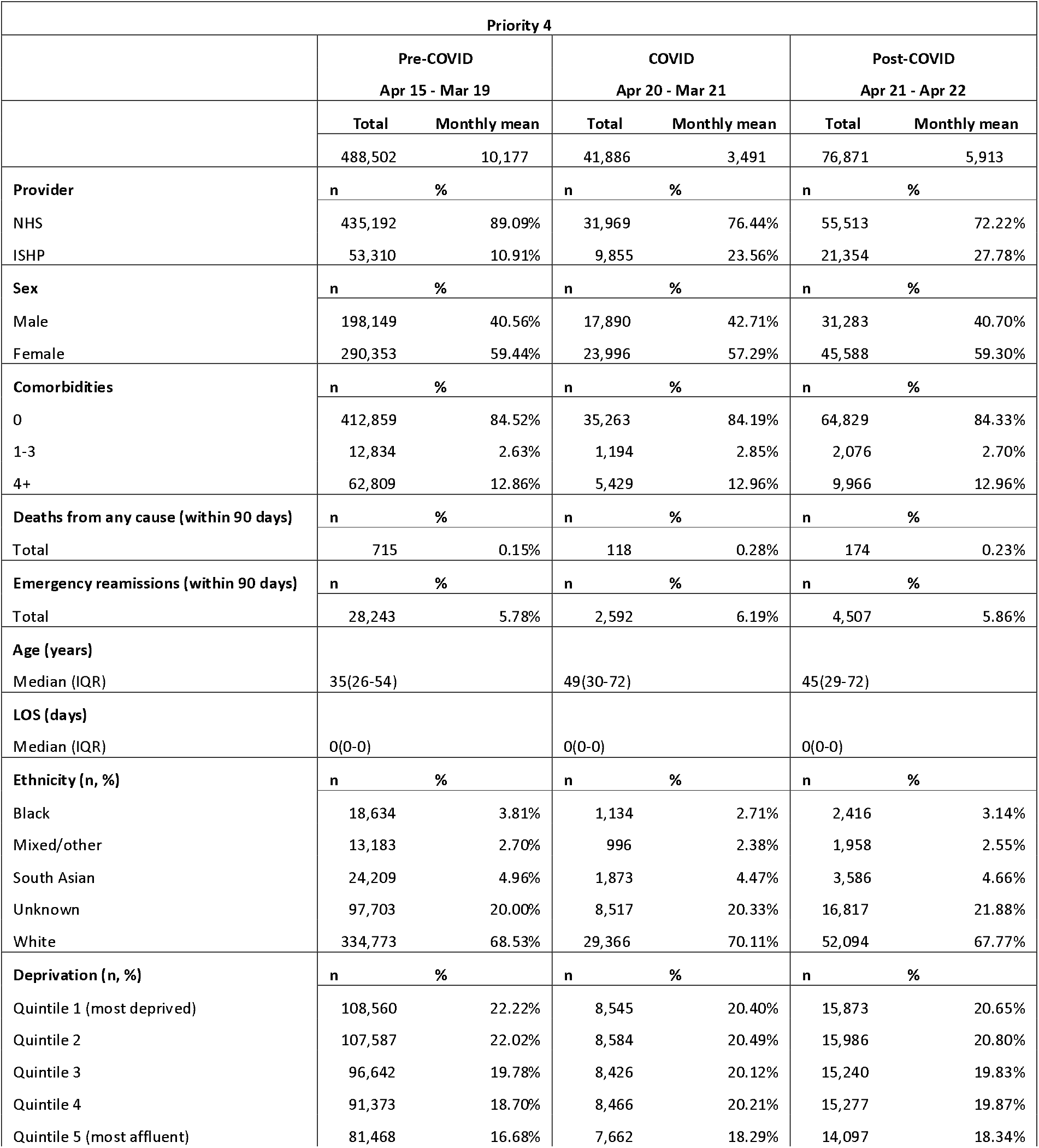
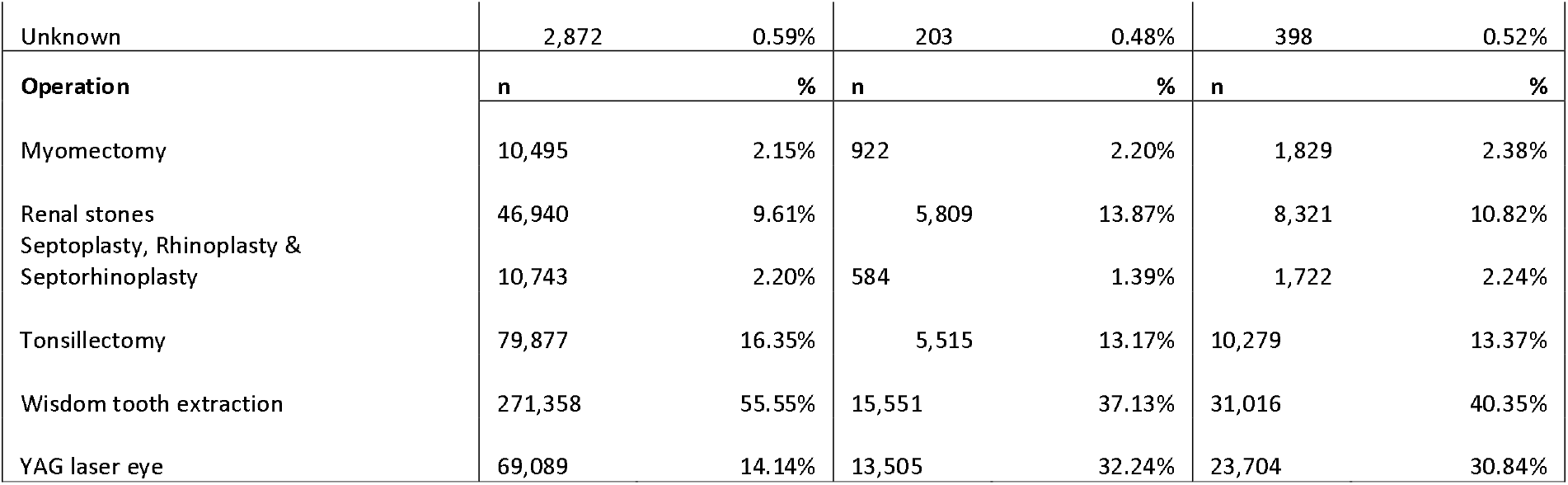
Total operations and outcomes for Priority 4

**Figure 2:**
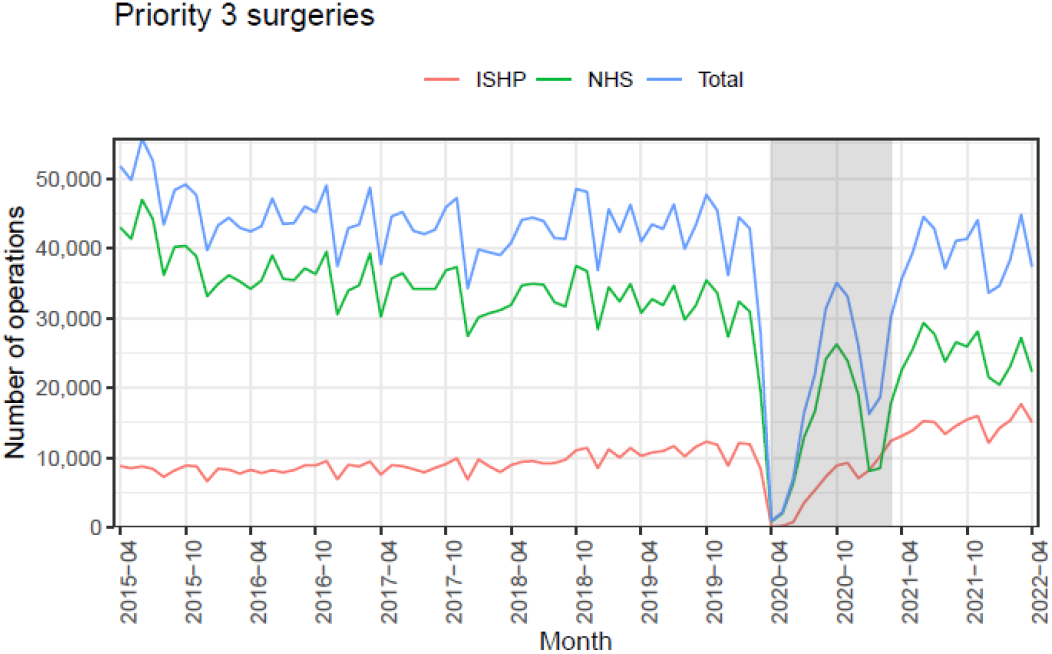
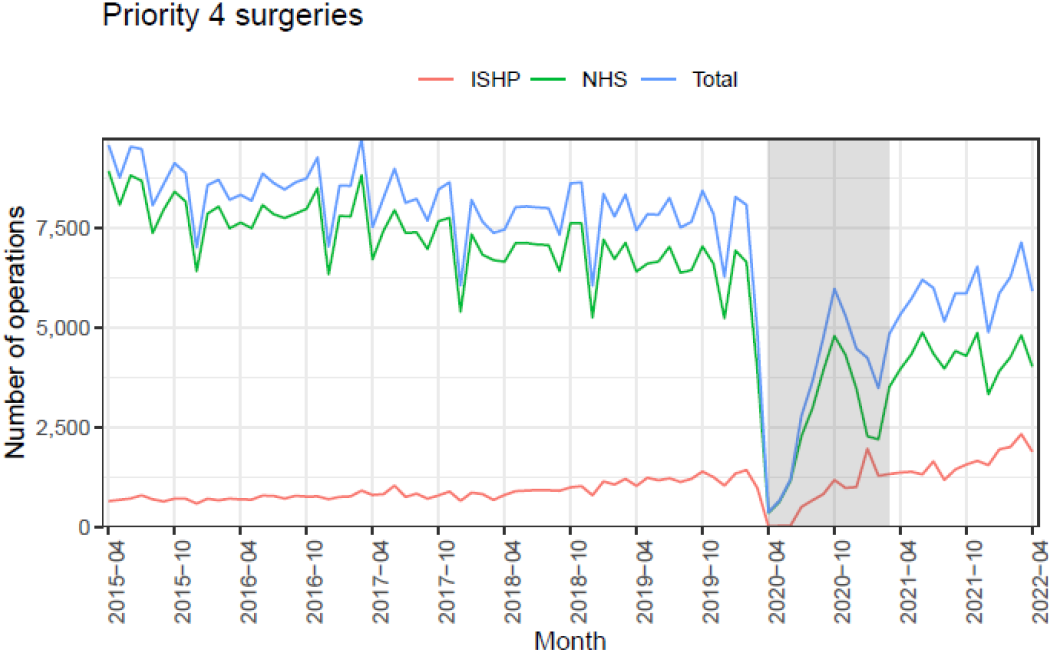
Operation numbers in NHS and ISHP hospitals over time and by priority ranking.

### Time-trends

Figure 2 shows the changing number of Priority 3 and 4, NHS-funded operations taking place in ISHPs and NHS hospitals over the three time windows. The data track the massive drop off in operations at the high points of the pandemic, followed by incomplete recovery.

### Proportions Treated in NHS vs Independent Hospitals Over Time-Windows

In both priority groups, we can see that there is a shift between operations in NHS hospitals vs independent sector hospitals in the pre- and post-COVID time windows. In Priority 3, 79% of operations were carried out by NHS providers before COVID and this falls to 63% post-COVID. Similarly, the percentage falls from 89% to 72% for Priority 4 operations.

Figure 3 shows the same operation numbers indexed to April 2015. These plots show that independent hospitals more than recovered after COVID and NHS hospitals are yet to return to pre-COVID levels. For Priority 3 operations, there were twice as many in independent hospitals at the start of 2022 as there had been in April 2015 and over three times as many for Priority 4. In NHS hospitals, however, there were half as many for Priority 3 and 4 as there had been in April 2015. This is in the direction hypothesised in the introduction (Question 1).

**Figure 3:**
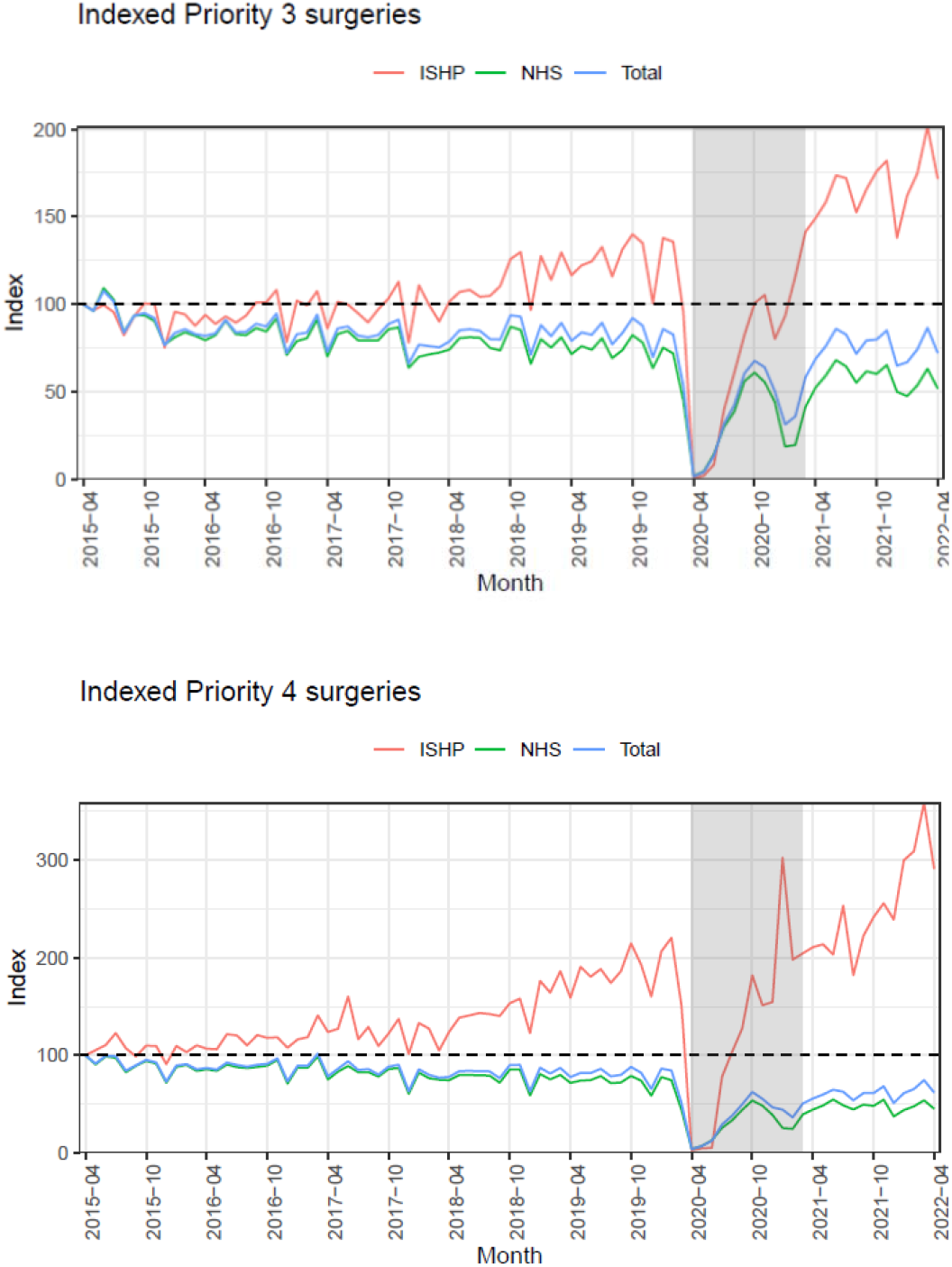
Indexed Operation numbers in NHS and ISHP hospitals over time and by priority ranking.

#### Prioritisation of Operation Types

Visual inspection of Figures 2 and 3 suggest that the drop in operation rates pre-COVID to post-COVID is of larger magnitude for priority 4 operations then for priority 3 operations. As stated earlier, the proportion of operations in the independent section is much higher for group 4 than for group 3. This is in the direction hypothesised under Question 3 in the introduction.

Table 4 shows the mean monthly percentage change for all operation types from pre-COVID to post-COVID. It is clear that the number of operations reduced for almost all operations types, except for YAG laser eye surgery.

**Table 4:** Mean monthly differences between pre and post COVID time windows in number of operations

|  | Mean monthly change | Mean monthly % change |
| --- | --- | --- |
| Priority 3 |  |  |
| All | -227.60 | 6.24% |
| Arthroplasty of joint | -5.39 | 11.64% |
| Lens replacement (cataracts) | -127.75 | 15.01% |
| Total cholecystectomy | -19.50 | 6.85% |
| Hernia repair | -7.48 | 12.79% |
| Hip replacement | -21.13 | 18.38% |
| Total abdominal hysterectomy | -4.86 | 1.05% |
| Knee replacement | -35.00 | 52.66% |
| Endoscopic resection of prostate | -6.50 | 4.66% |
| Priority 4 |  |  |
| All | -58.88 | 3.48% |
| Septoplasty, Rhinoplasty & Septorhinoplasty | -1.36 | 11.85% |
| Myomectomy | -0.71 | 9.39% |
| Renal stones | -12.56 | 0.35% |
| Tonsillectomy | -10.23 | 3.21% |
| Wisdom tooth extraction | -42.71 | 5.81% |
| YAG laser eye | 8.70 | 13.60% |

#### Effect of Socio-Economic Level and Ethnic Group

Supplementary Figure S1 shows surgery by deprivation quintile. There is no obvious visual trend by deprivation quintile. Supplementary Figure S2 shows operation numbers by ethnic group for Priority 3 surgeries in NHS hospitals and ISHPs. The same data are presented for Priority 4 surgeries in Supplementary Figure S3. The most noticeable feature is the spike in Priority 4 surgeries among white patients in ISHPs specifically. The spike referred to in Figure 6 seems to be driven by one operation only – YAG laser eye surgery.

#### Outcomes

The death rate among Priority 3 operations is higher in the COVID period by 0.07%, despite a lower proportion of people with co-morbidities in the COVID time window (Table 3). Nevertheless, there is a slight (0.1%) reduction in emergency readmissions in the COVID period. Deaths and readmissions are both increased in the COVID time window but, in this case, the proportion with co-morbidities increases slightly. The death rate for Priority 4 operations is also higher during COVID than pre-COVID but the comorbidity group proportions remain unchanged.

## DISCUSSION

### Key results

We found that, overall, operation rates have not recovered to their pre-COVID level. This has obvious implications for the reduction of waiting times and health effects for the population. Our data are silent on the reasons for failure for surgery rates to recover. Conceivably, this is simply part of a secular trend and some support for this idea comes from the trend line observed in Figure 2. Alternatively, and we think more plausibly, the COVID pandemic has done lasting structural damage that include staff shortages observed elsewhere in the economy.(5, 6)

It is clear that the independent sector has absorbed some of the increased demand for elective surgical services. Elsewhere, we have shown that outcomes for NHS-funded patients are better following operations in the independent sector compared to NHS hospitals after propensity score matching and across different risk levels.(7)

We find clear evidence that under conditions of high demand, higher value procedures are awarded increasing priority. Whether this trend has gone far enough is a topic worthy of further study, but it is reasonable to suppose that as the number of these procedures increases, so the cases selected are those which are causing the patient greater distress. Insofar as this is true, the utility gain per operation will rise as the numbers of such operations performed declines – the formal argument is laid out elsewhere.(8)

With regard to socioeconomic and ethnic groups, comparisons must be interpreted with caution because we do not know how denominators may be shifting in the population. In the particular issue of ethnicity, the proportions change because of rapidly changing willingness to declare an ethnic group (Tables 2 and 3). However, there is one signal that stands out from noise and that cannot plausibly be attributed to shifts in underlying proportions in the population – the spike in YAG eye operations. These are done mainly in White people and in independent hospitals. This enigmatic finding warrants further explanation.

### Limitations

There are always limitations when working with routinely collected healthcare data, mainly that they are not collected for the purpose of audit, service evaluation or research. There are possible coding errors and missing data, however our analysis is purely descriptive and shows the trends over a very large number of operations.

### Interpretation

The English NHS is responding to the crisis of rising waiting times by intervening at the supply side and the demand side. At the demand side, the government is producing an app that can direct patients to providers with the lowest waiting lists.(9) A possible problem with this policy is that it assumes that supply can be rapidly expanded to meet increased demand in organisations with currently low waiting times. At the supply side, the government is promoting High Volume Low Complexity Hubs, whose resources are ring-fenced to provide separate packages for elective operations in elective facilities.(10) A call has been announced by the National Institute for Health and Care Research (NIHR) to conduct a thorough investigation of these Hubs. In due course we will have evidence on whether Hubs can reduce waiting times overall. In the meantime, it would appear that the independent sector is absorbing some of the load of elective surgery, but with less discrimination between more and less urgent cases.

### Generalisability

The data used for this study are for the most frequently performed operations in each priority group. Therefore, our conclusions are strictly generalisable across these operation types.

## Data Availability

The protocol can be shared on request. Raw data and programming code is not available to share due to data sharing agreements with NHS Digital.

## OTHER INFORMATION

### Funding

This study was funded by the National Institute for Health and Care Research (NIHR) Applied Research Collaboration (ARC) West Midlands and the NIHR ARC East Midlands. Views expressed here are not necessarily those of any funder, the NHS, or the Department of Health and Social Care.

### Ethics

This database study was registered with the local Clinical Audit Department. The University of Birmingham Science, Technology, Engineering and Mathematics Ethical Review Committee board ruled that ethical approval could be waived and patient consent was not needed. Data were used in line with the data sharing agreement with NHS Digital.

## Notes

### Competing Interest Statement

The authors have declared no competing interest.

