## Supplementary for "Elective Surgery Before, During and After the COVID-19 Pandemic in England 2015 - 2022: A Database Study"

### Supplementary material

Supplementary Table S1: Surgical prioritisation criteria:

| 1a | Emergency procedures to be performed in less than 24 hours. |
| --- | --- |
| 1b | Procedures to be performed in less than 72 hours. |
| 2 | Procedures to be performed in less than 1 months. |
| 3 | Procedures to be performed in less than 3 months. |
| 4 | Procedures to be performed in over 3 months. |

*Supplementary Table S2: Surgeries and Codes*

| **Priority 3** | | |
| --- | --- | --- |
| Arthroplasty of joint | V203 | Intra-articular arthroplasty of temporomandibular joint |
|  | W491 | Primary prosthetic replacement of head of humerus using cement |
|  | W494 | Resurfacing hemiarthroplasty of head of humerus using cement |
|  | W500 | Conversion from previous uncemented prosthetic replacement of head of humerus |
|  | W501 | Primary prosthetic replacement of head of humerus not using cement |
|  | W502 | Conversion to prosthetic replacement of head of humerus not using cement |
|  | W503 | Revision of prosthetic replacement of head of humerus not using cement |
|  | W504 | Resurfacing hemiarthroplasty of head of humerus not using cement |
|  | W508 | Other specified prosthetic replacement of head of humerus not using cement |
|  | W509 | Unspecified prosthetic replacement of head of humerus not using cement |
|  | W510 | Conversion from previous prosthetic replacement of head of humerus NEC |
|  | W511 | Primary prosthetic replacement of head of humerus NEC |
|  | W512 | Conversion to prosthetic replacement of head of humerus NEC |
|  | W513 | Revision of prosthetic replacement of head of humerus NEC |
|  | W514 | Attention to prosthetic replacement of head of humerus NEC |
|  | W515 | Resurfacing hemiarthroplasty of head of humerus NEC |
|  | W518 | Other specified other prosthetic replacement of head of humerus |
|  | W519 | Unspecified other prosthetic replacement of head of humerus |
|  | W550 | Conversion from previous prosthetic interposition arthroplasty of joint |
|  | W551 | Primary prosthetic interposition arthroplasty of joint |
|  | W552 | Revision of prosthetic interposition arthroplasty of joint |
|  | W553 | Conversion to prosthetic interposition arthroplasty of joint |
|  | W554 | Attention to prosthetic interposition arthroplasty of joint NEC |
|  | W558 | Other specified prosthetic interposition reconstruction of joint |
|  | W559 | Unspecified prosthetic interposition reconstruction of joint |
|  | W560 | Conversion from previous interposition arthroplasty of joint NEC |
|  | W561 | Primary interposition arthroplasty of metatarsophalangeal joint NEC |
|  | W562 | Primary interposition arthroplasty of joint NEC |
|  | W563 | Revision of interposition arthroplasty of joint NEC |
|  | W564 | Conversion to interposition arthroplasty of joint NEC |
|  | W568 | Other specified other interposition reconstruction of joint |
|  | W569 | Unspecified other interposition reconstruction of joint |
|  | W570 | Conversion from previous excision arthroplasty of joint |
|  | W571 | Primary excision arthroplasty of first metatarsophalangeal joint |
|  | W572 | Primary excision arthroplasty of joint NEC |
|  | W573 | Revision of excision arthroplasty of joint |
|  | W574 | Conversion to excision arthroplasty of joint |
|  | W578 | Other specified excision reconstruction of joint |
|  | W579 | Unspecified excision reconstruction of joint |
|  | W580 | Conversion from previous resurfacing arthroplasty of joint |
|  | W581 | Primary resurfacing arthroplasty of joint |
|  | W582 | Revision of resurfacing arthroplasty of joint |
|  | W588 | Other specified other reconstruction of joint |
|  | W589 | Unspecified other reconstruction of joint |
| Total cholecystectomy | J183 | Total cholecystectomy NEC |
| Lens replacement (cataracts) | C751 | Insertion of prosthetic replacement for lens NEC |
|  | C754 | Insertion of prosthetic replacement for lens using suture fixation |
|  | C758 | Other specified prosthesis of lens |
|  | C759 | Unspecified prosthesis of lens |
| Hernia repair | T212 | Repair of recurrent inguinal hernia using insert of prosthetic material |
|  | T242 | Repair of umbilical hernia using insert of prosthetic material |
|  | T243 | Repair of umbilical hernia using sutures |
|  | T272 | Repair of ventral hernia using insert of prosthetic material |
| Hip replacement | W371 | Primary total prosthetic replacement of hip joint using cement |
|  | W381 | Primary total prosthetic replacement of hip joint not using cement |
|  | W391 | Primary total prosthetic replacement of hip joint NEC |
|  | W931 | Primary hybrid prosthetic replacement of hip joint using cemented acetabular component |
|  | W941 | Primary hybrid prosthetic replacement of hip joint using cemented femoral component |
| Total abdominal hysterectomy | Q074 | Total abdominal hysterectomy NEC |
| Knee replacement | W401 | Primary total prosthetic replacement of knee joint using cement |
|  | W411 | Primary total prosthetic replacement of knee joint not using cement |
|  | W421 | Primary total prosthetic replacement of knee joint NEC |
| Endoscopic resection of prostate | M653 | Endoscopic resection of prostate NEC |
| **Priority 4** | | |
| Wisdom tooth extraction | F091 | Surgical removal of impacted wisdom tooth |
|  | F093 | Surgical removal of wisdom tooth NEC |
| Tonsillectomy | F341 | Bilateral dissection tonsillectomy |
|  | F342 | Bilateral guillotine tonsillectomy |
|  | F343 | Bilateral laser tonsillectomy |
|  | F344 | Bilateral excision of tonsil NEC |
|  | F345 | Excision of remnant of tonsil |
|  | F346 | Excision of lingual tonsil |
|  | F347 | Bilateral coblation tonsillectomy |
|  | F348 | Other specified excision of tonsil |
|  | F349 | Unspecified excision of tonsil |
| Septoplasty, Rhinoplasty & Septorhinoplasty | E023 | Septorhinoplasty using implant |
|  | E024 | Septorhinoplasty using graft |
|  | E025 | Reduction rhinoplasty |
|  | E026 | Rhinoplasty NEC |
|  | E028 | Other specified plastic operations on nose |
|  | E174 | Lateral rhinotomy into nasal sinus NEC |
| Myomectomy | Q092 | Open myomectomy |
| Renal stones | M141 | Extracorporeal shock wave lithotripsy of calculus of kidney |
|  | M148 | Other specified extracorporeal fragmentation of calculus of kidney |
|  | M149 | Unspecified extracorporeal fragmentation of calculus of kidney |
|  | M311 | Extracorporeal shockwave lithotripsy of calculus of ureter |
| Tonsillectomy | F341 | Bilateral dissection tonsillectomy |
|  | F342 | Bilateral guillotine tonsillectomy |
|  | F343 | Bilateral laser tonsillectomy |
|  | F344 | Bilateral excision of tonsil NEC |
|  | F345 | Excision of remnant of tonsil |
|  | F346 | Excision of lingual tonsil |
|  | F347 | Bilateral coblation tonsillectomy |
|  | F348 | Other specified excision of tonsil |
|  | F349 | Unspecified excision of tonsil |
| Wisdom tooth extraction | F091 | Surgical removal of impacted wisdom tooth |
|  | F093 | Surgical removal of wisdom tooth NEC |
| YAG laser eye | C732 | Capsulotomy of anterior lens capsule |
|  | C733 | Capsulotomy of posterior lens capsule |
|  | C734 | Capsulotomy of lens NEC |

*Supplementary Figure S1: Operation numbers by deprivation quintile in NHS and ISHP providers*

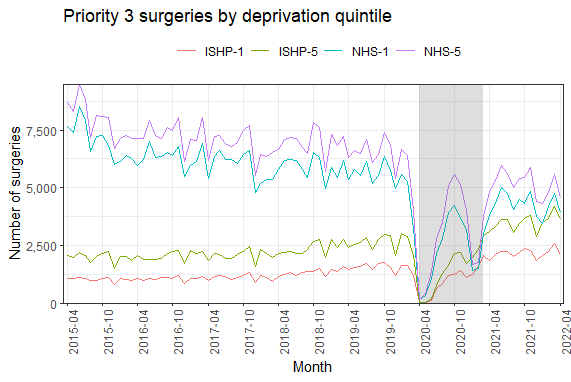

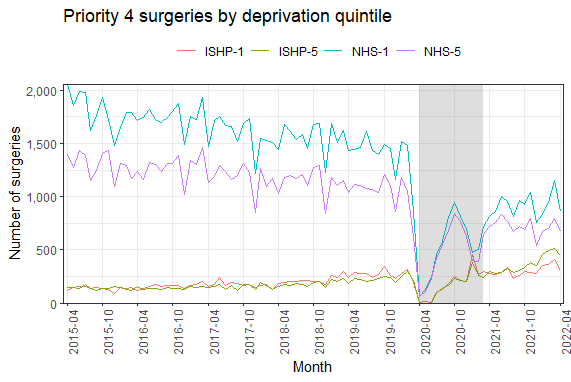

*Supplementary Figure S2: Operation numbers by ethnic group for Priority 3 patients*

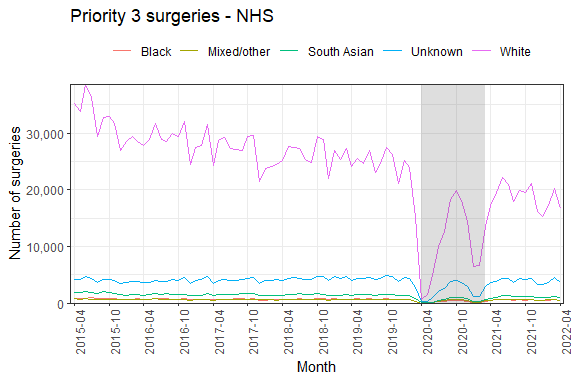

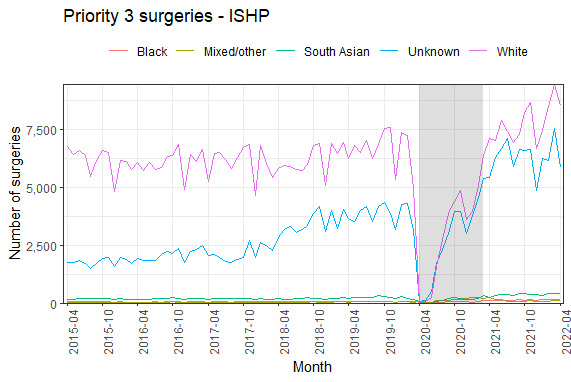

*Supplementary Figure S3: Operation numbers by ethnic group for Priority 4 patients*

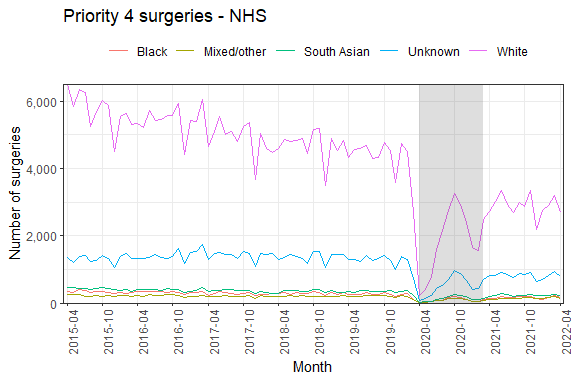

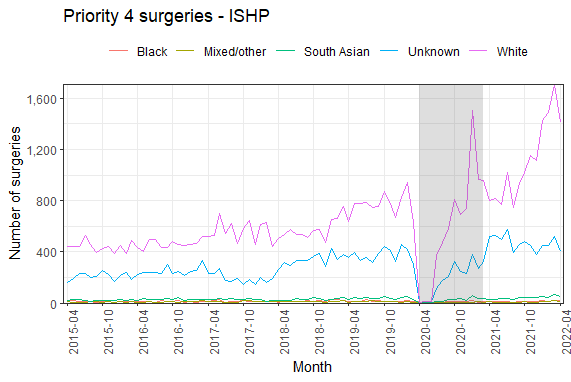
